# A comparison of sleep metrics from mid-thigh and low-back accelerometers to wrist based data using open-source algorithms

**DOI:** 10.1101/2024.11.10.24317079

**Authors:** Geoffrey Passfield, Lisa Mackay, Catherine Crofts, Grant Schofield

## Abstract

**Introduction:** Wearable accelerometers are a valuable tool for monitoring sleep, sedentary behaviour, and physical activity patterns within 24h time-use in free-living environments. While wrist-worn accelerometers are favoured for monitoring sleep, they do not accurately distinguish between sitting and lying positions (Narayanan et al., 2020). This study aims to determine whether back or thigh-mounted accelerometers yield sleep metrics comparable to wrist-worn devices using an open-source algorithm originally validated for the wrist.

**Methods:** Data from 20 healthy sleepers were collected using Axivity AX3 accelerometers. Participants wore accelerometers on their right thigh, low-back, and wrist for one night of sleep in their own bed. Sleep metrics were calculated using the van Hees algorithm through the GGIR package in R. The primary outcomes were: Total Sleep Time (TST), Wake After Sleep Onset (WASO), Awakenings (AWK), Sleep Efficiency (SE), Sleep Interval (SI) and Sleep Onset Timestamp (SOT). Within-subject ANOVA with Tukey’s post hoc, Pearson correlation coefficients, Bland-Altman plots, and Cohen’s d were used to assess the comparability of sleep metrics between the body placements.

**Results:** Data analysis included all 20 participants. Mid-thigh accelerometers demonstrated a strong linear relationship with wrist accelerometers across all metrics (*r* = 0.86-0.98). Bland-Altman plots demonstrated a narrow 95% confidence interval suggesting that wrist and mid-thigh metrics are in good agreement, except for AWK which is slightly underestimated by the mid-thigh device. Conversely, low-back accelerometers demonstrated moderate linear relationship with the wrist (*r* = 0.63-0.98) and the Bland-Altman results showed wide limits of agreement with significant overestimations of TST, SE, SI and underestimations of WASO, AWK, SOT. Cohen’s d demonstrated small differences between mid-thigh and wrist devices, except for AWK (d= 0.42). Low-back values for WASO, SE, and AWK showed moderate differences.

**Conclusions:** This analysis demonstrates that the mid-thigh accelerometer yields comparable sleep metrics to wrist-worn devices when processed with the van Hees algorithm.

## Introduction

The emerging field of 24-hour behavioural time use epidemiology is calling for valid and reliable measures of behavioural sleep, physical activity, and sedentary behaviour to better understand the relationship between the 24-hour composition of behaviour and health.

Understanding the interdependence between different facets of human function is essential in health research, as it enables us to measure their influences effectively and comprehend how they relate to overall health (Crowley et al., 2019; Johansson et al., 2022; Perez-Pozuelo et al., 2020). Therefore, there is a need to ensure that these measures can reliably and accurately measure all three behaviours. Finding a common location that provides valid, useful results for both wake and sleep phases of an individual’s day is compelling as an endeavour as it will help to simplify the research process. Additionally, it may also unlock further insights from retrospective analysis for many studies using data that has already been collected.

Sedentary, light, and moderate to vigorous physical behaviours are collectively considered to encompass all waking behaviours with sleep consuming the remaining time in a 24-hour period (Rosenberger, 2019). The time spent in each aspect of behaviour throughout the day is mutually exclusive and perfectly collinear. In other words, you can only be performing one of these behaviours at a time, and the sum time spent in each behaviour, will result in a total of 24 hours (Pedišić, 2014). The distribution of these component behaviours has been associated to health outcomes related to both physical and mental health (Baillot, 2022; Brakenridge et al., 2021; Dumuid et al., 2022). Of these behaviours, sleep is encouraged to take around one-third of the 24-hour period and is also in and of itself highly associated with health outcomes (Cappuccio et al., 2010; Watson et al., 2015). It is vital in the recovery from previous waking events and in the preparation of the body and mind for subsequent waking activities. Ineffective and/or insufficient sleep has been associated with both immediate and long term health outcome risks including premature mortality, cardiovascular disease, hypertension, inflammation, obesity, diabetes and impaired glucose tolerance, and psychiatric disorders, such as anxiety and depression (Ferrie et al., 2011). Insufficient sleep in the general population has been identified by countries around the globe and along with the personal health risk factors go with it economic costs to personal finance, business, and government (Hafner, 2017). Indeed, sleep is a public health issue that is worthy of investigation.

Sleep tracking using wearable accelerometers has gained substantial attention as a non-invasive and objective method to monitor sleep patterns and behaviours in free-living environments. The use of accelerometers offers the advantage of capturing continuous data over extended periods, providing insights into sleep quantity and quality, as well as its relationships with movement behaviours across a full 24-hour period. As such the use of wearable accelerometers over 24-hour periods and longer is increasingly popular as an objective research approach for tracking both waking and sleep behaviours (Hills et al., 2014; Sundararajan et al., 2021). The ways in which accelerometric data can be interpreted by these devices is dependent on factors including body placement, sampling frequency, and computational methods.

The most common body placement locations used for accelerometery studies include the wrist, hip, and thigh (Evenson et al., 2022; Lettink et al., 2022). Gao et al. (2021) identified a shift from hip to wrist worn accelerometers citing advantages of improved compliance, being less intrusive, improved capability of assessing sleep behaviours, along with the capability of capturing more upper extremity movements. Nonetheless, this approach has limitations in accurately detecting body posture (Rosenberger, 2019). A wrist mounted device has no ability to determine whether a person is standing, sitting, or lying, a critical aspect in understanding sleep behaviour or sedentary behaviour. The definition of sedentary behaviour requires that the individual be waking as well as sitting or lying (Tremblay et al., 2017). Thus, differentiating between standing, and sitting or lying is necessary for achieving accurate measurements of different waking physical behaviours. Stewart et al. (2018) found that a dual accelerometer system that placed devices on the mid-thigh and low back performed well in identifying the positions and activities of standing, sitting, lying, walking, and running. Narayanan et al. (2020) furthered this, validating its application in semi free-living conditions. However, it was also noted that this work would need expansion in its measurement of other components of behaviour such as sleep. As such more work needs to be completed to validate device placements and data processing algorithms to measure physical activity, sedentary behaviour, and sleep concurrently.

Sampling frequency in data collection is another crucial consideration that can influence the granularity and accuracy of sleep and physical behaviour tracking outcomes. The sampling frequency, also known as sampling rate, refers to the number of data points taken by the accelerometer each second. A higher sampling frequency translates to more data points captured within a given timeframe. Higher sampling rates enable a more accurate representation of movements during both wake and sleep phases. The time a device can record for is also dependent on the sampling frequency as higher sampling rates will reduce battery length. A balance must therefore be decided upon where the researcher is able to record the highest detail while still ensuring that the full duration of the recording period is completed. Migueles et al. (2017) suggests that using the highest frequency possible is the recommended approach to sampling. They go on to suggest that 100Hz is preferable over frequency multiples of 30Hz when researchers are filtering and processing the signal on their own (e.g., open-source software). The Axivity brand AX3 is a commonly used accelerometer in sleep and 24-hour time use research. It has a sampling frequency that can be specified from 12.5 to 3200Hz with the default setting being 100Hz and can be used with open-source software. AX3 devices set to record at 100Hz are reported by Axivity (Axivity operating manual), to have a battery life of 14 days of continuous recording. As such we can see a need to have a system of sleep metric analysis that uses the default setting for AX3 accelerometers.

To interpret the raw data collected from any accelerometric recording device a computer algorithm is required to synthesise the enormous volume of data points. The computational algorithms used to analyse accelerometery data play a pivotal role in translating raw data into meaningful metrics, for example, sleep duration or minutes of moderate physical activity. In the endeavour to create reproducible, comparable, and composite insights that accurately reflect habits of 24-hour time use, a key component of the task is using methods that are transparent. Proprietary software, while common, brings challenges like transparency and consistency in algorithmic computation (Karas et al., 2019). Non-proprietary and open-source methods offer several benefits over proprietary offerings including transparency, the avoidance of vendor lock-in, long-term availability, and cost efficiency. These aspects all contribute to the promotion of collaboration and reproducibility. It is essential for research purposes that raw frequencies are collected and disclosed, and that non-proprietary, open-source algorithms are able to be shared and applied. The emergence of open-source algorithms have paved the way for researchers to compare results across samples in a transparent and innovative manner. These open-source algorithms also have the advantage of being interchangeable with most popular brands of accelerometery recorders like Axivity and ActiGraph. Indeed, Crowley et al. (2019) found that these brands were interchangeable in their ability to report accelerometric data when using the same algorithm for calculation of metrics.

This study aims to address the above intricacies by investigating the validation of mid-thigh, and low-back mounted accelerometers against a computational algorithm primarily designed for wrist-mounted accelerometery where the sampling frequency is set to 100Hz. The primary objective is to assess whether either the mid-thigh or low-back placements yield comparable and reliable sleep metrics, when compared against wrist data. By doing so, this research seeks to contribute to the advancement of accurate and comprehensive sleep tracking methodologies that can be integrated into 24-hour time use research and will also allow retrospective analysis for many studies.

## Methods

Participants were recruited from the general public via an advertisement and word of mouth. The inclusion criteria were: at least 20 years of age and self-reported healthy sleepers. For the purposes of this study, healthy sleepers were operationally defined as individuals who did not utilise or necessitate sleep aids, thereby ensuring the absence of substances or devices that could impede or constrain normal sleep function or physical mobility. All participants provided written informed consent. Ethical approval was obtained from Auckland University of Technology ethics committee (AUTEC 22/300).

### Data Collection

After providing signed informed consent, participants completed a sleep diary and wore three Axivity AX3 accelerometers simultaneously for a single night of sleep at home in a free-living environment.

### Accelerometers

The Axivity AX3 device is a small, lightweight, and waterproof accelerometer (23 x 32.5 x 8.9 mm; 11 g) that can be affixed using medical tape, adhesive pouches, or worn on a wrist band. AX3 devices have been previously used and validated in research involving movement behaviour as well as sleep (Johansson et al., 2022; Narayanan et al., 2020; Plekhanova et al., 2020). Each Axivity AX3 accelerometer was pre-set to automatically start recording at 7pm until 11am the next day. Sample rate was set to 100 Hz with a sensitivity of ±8 *g*.

Participants placed the devices on their mid anterior thigh, lower-back offset from the spine, and posterior wrist, following the instructions provided. All devices were worn on the right side of the body (see Figure 1). The devices were affixed using a self-adhesive pouch (thigh, back) labelled with location and directionality instructions, or a wrist-watch style strap.

**Figure 1.**
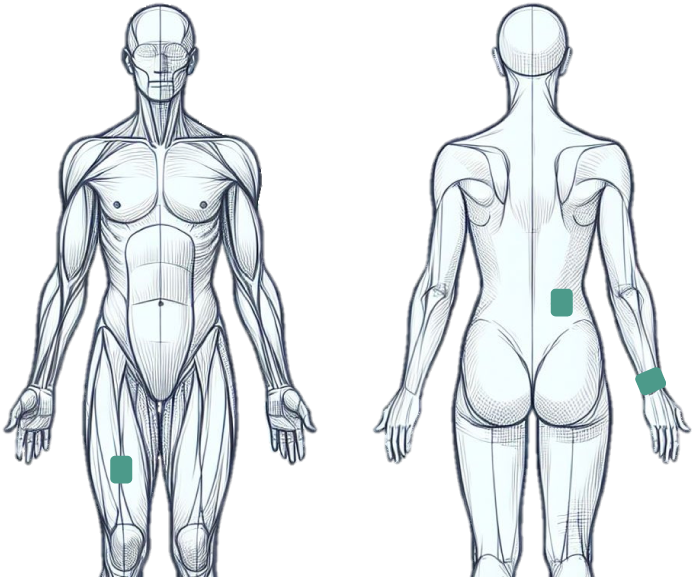
Placement locations for AX3 devices.

Participants were instructed to place the devices at 6pm and remove the device after they had gotten out of bed and completed their sleep diary the next day.

### Sleep Diary & Data Processing

On a paper-based sleep diary, participants recorded the time they attached and removed the device, the time they got into bed, the time they tried to go to sleep, the approximate time they woke up (final awakening time), and the time they got up.

Using OMGUI Open Movement software (version 1.0.0.30; Open Movement, Newcastle University, UK), raw accelerometer data was downloaded and logged into ID folders for processing. The Open-source software ‘R package’ (R Core Team, 2021) was used to process the accelerometric data for all device placements. The software package GGIR (Generalized Graded Intensity Model R-package) (Migueles et al., 2019) was used to synthesize the data into sleep metrics. The van Hees et al. (2015) algorithm was chosen for this study from the four different algorithms offered by GGIR for measuring sleep metrics. Unlike earlier algorithms that relied on acceleration magnitude for movement/sleep detection (Cole et al., 1992; Galland, 2012; Sadeh, 1994), the van Hees algorithm interprets sleep through body kinematics using (arm) angles and lack of movement to identify sleep and has also been validated against sleep tracking gold standard PSG.

The sleep diary data was used as a guider by the algorithm. As a guider, the sleep diary data did not provide a rigid time for the sleep algorithm to use. Instead, it directed the algorithm to the approximate start and end points of sleep measurements. The computed sleep data was then exported to Microsoft Excel as a .csv file. The derived sleep metrics for analysis were total sleep time (TST), wake after sleep onset (WASO), awakenings (AWK), sleep interval (SI), sleep efficiency (SE). Sleep onset timestamp (SOT) was also used as a device location comparator although it is not specifically a sleep metric.

### Analysis

Statistical analysis was performed in The jamovi project (2022) (Version 2.3.26) and statistical significance was set at α = 0.05. First, visual inspection was undertaken to initially assess whether further statistical investigation was worthwhile. A within-subjects analysis of variance (ANOVA) was conducted to compare mean estimates of sleep metrics across the three device placements (thigh, back, wrist). Post-hoc pairwise comparisons using Tukey’s test were performed to identify pairwise differences between sleep metrics. Cohen’s d was computed to interpret the effect size of the pairwise differences in means. Pearson correlation coefficients were calculated to assess the linear relationship between wrist-thigh and wrist-back estimates of sleep metrics. An *r* value of ≥ 0.8 was considered to be a level of high convergent validity (Mukaka, 2012). Finally, Bland-Altman plots were generated for wrist-thigh and wrist-back estimates of sleep metrics to assess the level of agreement between device placements.

## Results

A total of 20 individuals (40% male) aged 20-65 consented to and completed the single night study. No further demographic data were collected as the primary objective was to compare sleep metric data from different accelerometer placements, rather than provide generalisable estimates of sleep metrics.

Visual analysis of minute-by-minute within-participant sleep data (e.g., Figure 2) showed that there were large periods of sleep alignment across all device placement sites for all participants. Although not all wake periods aligned identically, overall, the alignment of sleep between body locations appeared to be good for each participant. During the sleep period, when wake events did not directly align between body locations, it was common, to observe similar lengths of wake time either slightly before or after. Large discrepancies were minimal but did occur. Three participants of the study recorded visually significant gaps in the mid-thigh and/or low-back devices in comparison to the wrist. One during the sleep period and two at the beginning of the sleep period.

**Figure 2.**
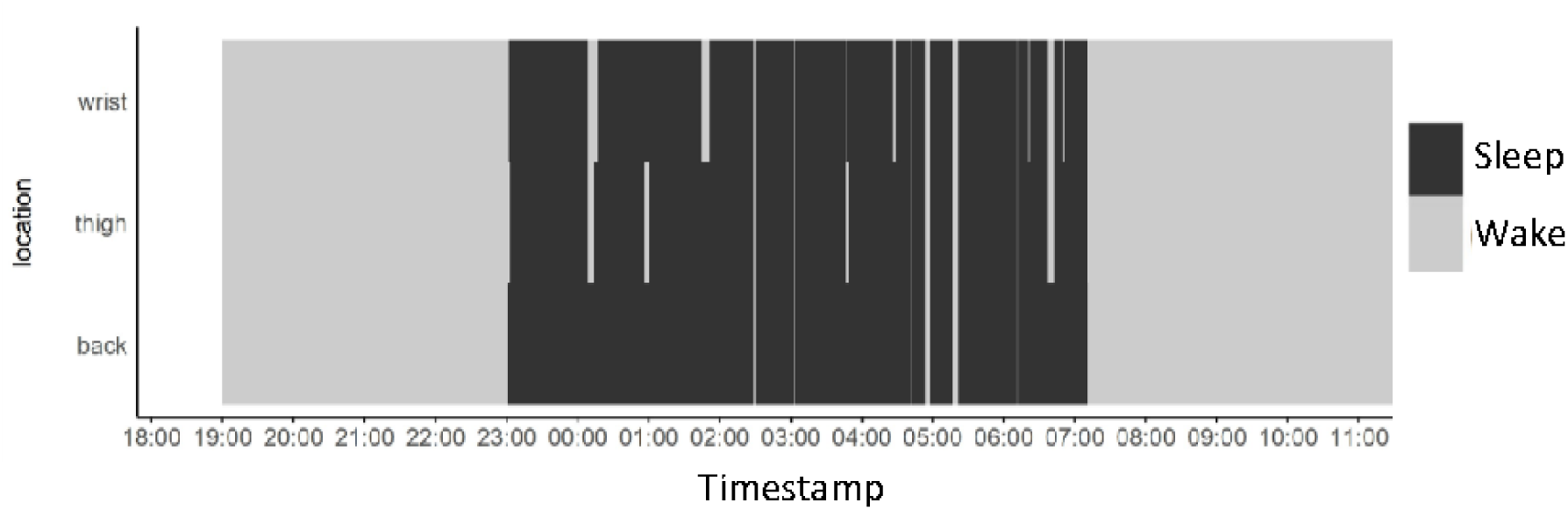
Example of visual sleep summary of each device per participant.

Mean values of the wrist group were less than the mid-thigh group for TST, SE and SI. Significant differences in mean value were identified in AWK for both the mid-thigh and low-back locations compared to the wrist. Low-back sleep metric mean values varied greater compared to the wrist. The range values for the low-back device however were generally smaller than both the wrist and mid-thigh groups.

Within-subjects ANOVA was performed to compare the sleep metrics determined by the van Hees wrist algorithm across the different device placements. There was a statistically significant difference between device placements in all sleep metrics (see Table 2) except for SOT (*F*(2, 38) = 3.14, *p* = 0.055). Tukey’s post-hoc analysis showed that most of the statistically significant differences were observed comparing the wrist to the low-back, and the low-back to the mid-thigh location. For the wrist-thigh comparison, only estimates of AWK were statistically different (*p* = .007), whereas for the wrist-back comparison, statistically significant differences were observed for TST, WASO, SE and AWK (*p* > .005).

**Table 1.** Descriptives for Sleep Metrics taken from the wrist, mid-thigh, and low-back body locations.

| Sleep metric | N | Wrist |  |  | Thigh |  |  | Back |  |  |
| --- | --- | --- | --- | --- | --- | --- | --- | --- | --- | --- |
|  |  | Mean | SD | Range | Mean | SD | Range | Mean | SD | Range |
| TST, min | 20 | 403.10 | 60.62 | 204.42 | 410.01 | 59.99 | 224.64 | 425.57 | 59.26 | 203.82 |
| WASO, min | 20 | 60.57 | 31.53 | 133.86 | 56.87 | 31.59 | 115.92 | 45.80 | 26.46 | 106.32 |
| AWK, n | 20 | 16.95 | 5.72 | 24.00 | 14.55 | 5.73 | 25.00 | 13.50 | 6.13 | 25.00 |
| SI, min | 20 | 463.68 | 68.54 | 284.75 | 466.88 | 65.14 | 262.91 | 471.38 | 63.11 | 264.33 |
| SE, % | 20 | 0.87 | 0.06 | 0.22 | 0.88 | 0.06 | 0.21 | 0.90 | 0.05 | 0.20 |

**Table 2.** Results of Repeated Measures ANOVA and Tuckey Post hoc analysis comparing sleep metrics from thigh, back and wrist device placements.

| Repeated measures ANOVA table |  |  |  |  |  | Tukey post hoc analysis |  |  |  |  |  |  |  |  |  |  |  |  |  |  |
| --- | --- | --- | --- | --- | --- | --- | --- | --- | --- | --- | --- | --- | --- | --- | --- | --- | --- | --- | --- | --- |
|  |  |  |  |  |  | Thigh vs. Wrist |  |  |  |  | Back vs. Wrist |  |  |  |  | Thigh vs. Back |  |  |  |  |
| Sleep Metric | SS | df | MS | F | p – value | Mean Difference | SE | df | t | p<sub>tukey</sub> | Mean Difference | SE | df | t | p<sub>tukey</sub> | Mean Difference | SE | df | t | p<sub>tukey</sub> |
| TST, min | 5297 | 2 | 2649 | 10.6 | < 0.001 | 6.91 | 4.58 | 19.0 | 1.51 | 0.309 | 22.47 | 5.23 | 19.0 | 4.30 | 0.001 | -15.56 | 5.16 | 19.0 | -3.02 | 0.019 |
| WASO, min | 2363 | 2 | 1181 | 6.96 | 0.003 | -3.70 | 3.70 | 19.0 | -1.00 | 0.586 | -14.77 | 4.19 | 19.0 | -3.53 | 0.006 | -11.07 | 4.44 | 19.0 | -2.50 | 0.055 |
| SE, % | 0.01 | 2 | 0.01 | 7.78 | 0.001 | 0.01 | 0.01 | 19.0 | 1.17 | 0.486 | 0.03 | 0.01 | 19.0 | 3.69 | 0.004 | -0.02 | 0.01 | 19.0 | -2.55 | 0.049 |
| AWK, count | 125 | 2 | 62.55 | 7.90 | 0.001 | -2.40 | 0.69 | 19.0 | -3.48 | 0.007 | -3.45 | 1.15 | 19.0 | -3.01 | 0.019 | -1.05 | 0.766 | 19.0 | -1.37 | 0.375 |
| SI, min | 598 | 2 | 299.2 | 3.39 | 0.044 | 3.20 | 3.36 | 19.0 | -0.95 | 0.614 | 7.70 | 3.18 | 19.0 | 2.42 | 0.064 | -4.50 | 2.25 | 19.0 | -1.997 | 0.140 |
| SOT difference, min | 378 | 2 | 189.2 | 3.14 | 0.055 | -3.04 | 2.74 | 19.0 | -1.11 | 0.521 | -6.15 | 2.69 | 19.0 | -2.28 | 0.083 | 3.11 | 1.82 | 19.0 | 1.71 | 0.226 |

Pearson correlation coefficients showed high to very high correlations between the wrist and mid-thigh placements. This strong correlation carried through for all sleep metrics with *r* values ranging from 0.86 to 0.98 with *p* < 0.001 (as shown in Table 3). The strongest correlation was SI, *r* =0.98 (*p* < 0.001) within the wrist-thigh comparison. WASO, SE, and AWK shared the lowest correlation value at *r* = 0.86 (p < 0.001) of wrist-thigh comparisons. Only two of the sleep metrics (AWK, *r* =0.63 (*p* < 0.003); SE, *r* =0.78 (*p* < 0.001)) did not reach the determined *r* value and were both found in wrist-low-back comparisons. AWK within the wrist-low-back comparison was the lowest of all the correlation values calculated.

**Table 3.** Results of Bland-Altman and Pearson Correlation analysis comparing sleep metrics obtained from mid-thigh and wrist device placements.

| N=20 | Mid-thigh compared to wrist |  |  |  |  |  | Pearson correlation, r | p value | 95% CI |
| --- | --- | --- | --- | --- | --- | --- | --- | --- | --- |
|  | Bias | 95% CI | Lower LoA | 95% CI | Upper LoA | 95% CI |  |  |  |
| TST, min | 6.91 | -2.68, 16.50 | -33.25 | -49.91, -16.6 | 47.06 | 30.40, 63.70 | 0.94 | p < 0.001 | 0.86, 0.98 |
| WASO, min | 3.70 | -11.50, 4.05 | -36.17 | -49.70, -22.70 | 28.76 | 15.30, 42.24 | 0.86 | p < 0.001 | 0.68, 0.94 |
| SE, % | 0.86 | -0.68, 2.40 | -5.59 | -8.27, -2.91 | 7.31 | 4.63, 9.98 | 0.86 | p < 0.001 | 0.68, 0.95 |
| AWK, n | -2.40 | -3.84, -0.96 | -8.45 | -10.96, -5.94 | 3.65 | 1.14, 6.16 | 0.86 | p < 0.001 | 0.66, 0.94 |
| SI, min | 3.20 | -3.82, 10.20 | -26.23 | -38.45, -14.00 | 32.64 | 20.42, 44.90 | 0.98 | p < 0.001 | 0.94, 0.99 |
| SOT difference, min | -3.04 | -8.78, 2.70 | -27.07 | -37.05, -17.10 | 21.00 | 11.02, 30.97 | 0.97 | p < 0.001 | 0.92, 0.99 |

Abbreviation: CI, confidence interval
LoA, Limits of Agreement

Bland-Altman plots showed that the thigh-mounted device slightly overestimated TST, SE, and SI, and slightly underestimated WASO, AWK, and SOT compared to the wrist-mounted device (Table 4). However, the 95% confidence intervals of the bias estimate included zero for all metrics except AWK, suggesting that only AWK was systematically underestimated by the thigh unit (95% CI -3.84, -0.96 awakenings per night). The widest limits of agreement (LoA) was observed for TST (-33.3 min to 47.1 min; range 80.3 min), while the LoA for SI was -26.2 min to 32.6 min (range 58.9 min). Figure 3 depicts the variation in sleep metrics between the two device placements, including the presence of outliers. For TST, there were two outliers of the LoA, whereas one outlier was observed for WASO, SE, SI and SOT. These outliers all came from the same four

**Figure 3.**
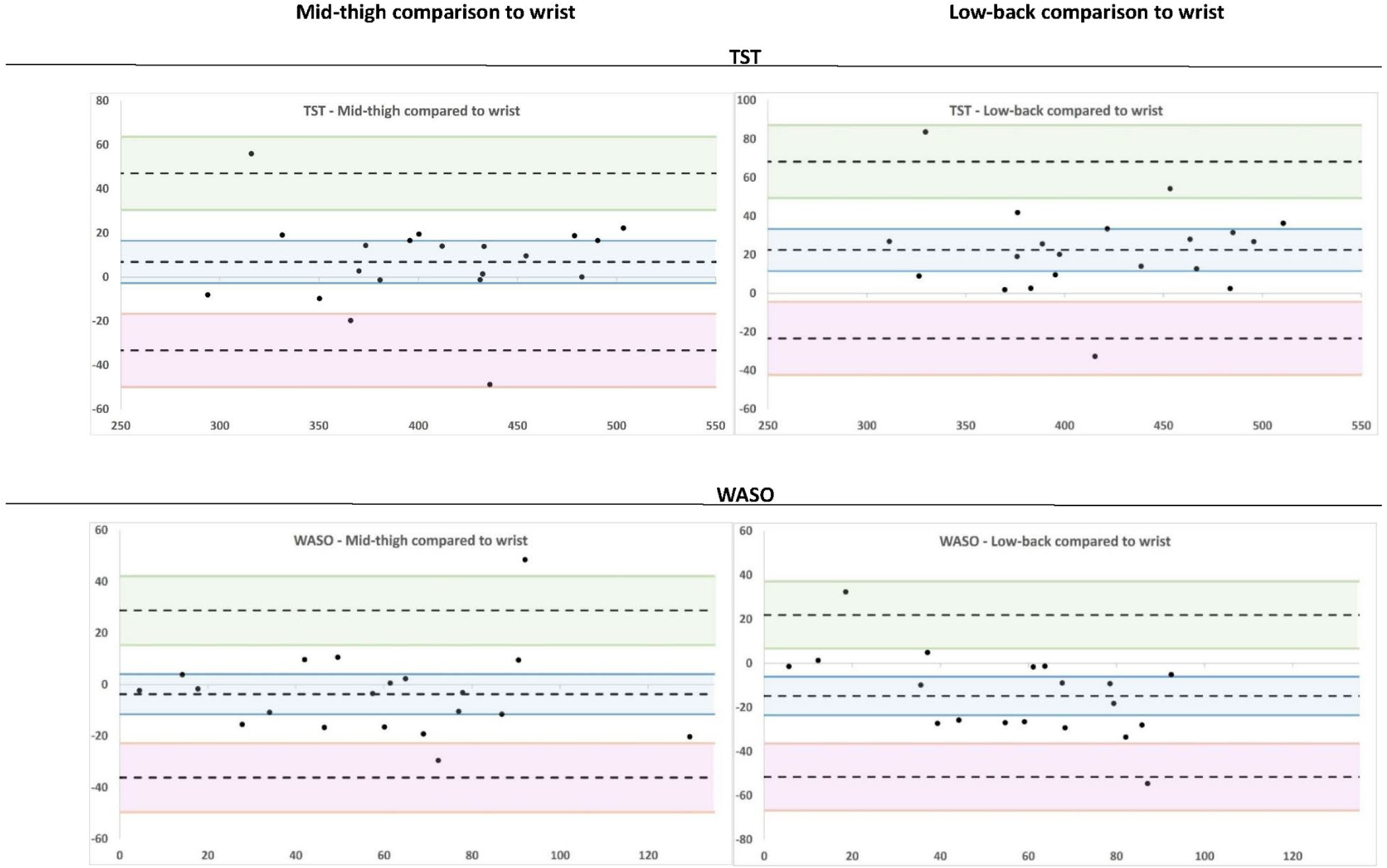
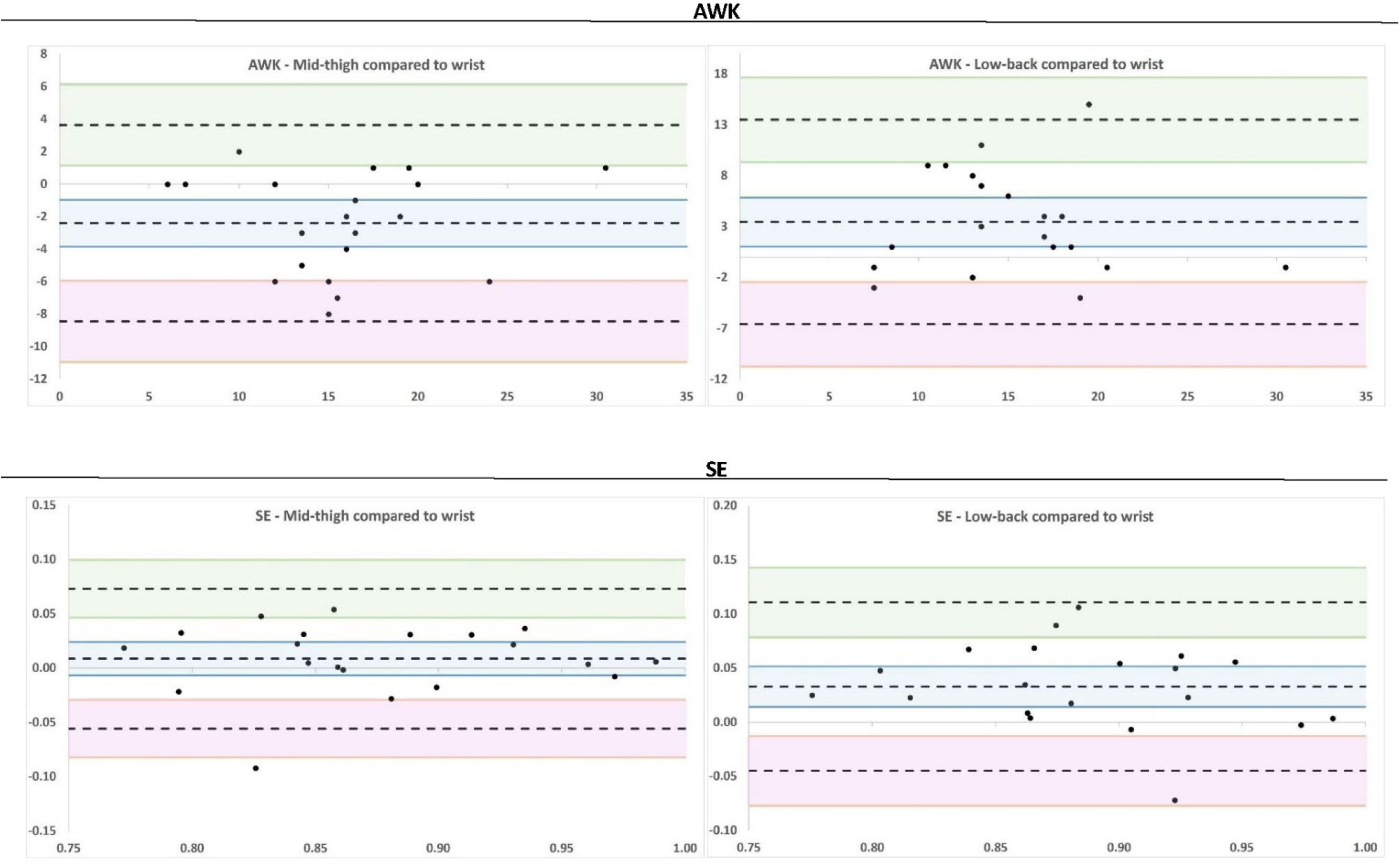
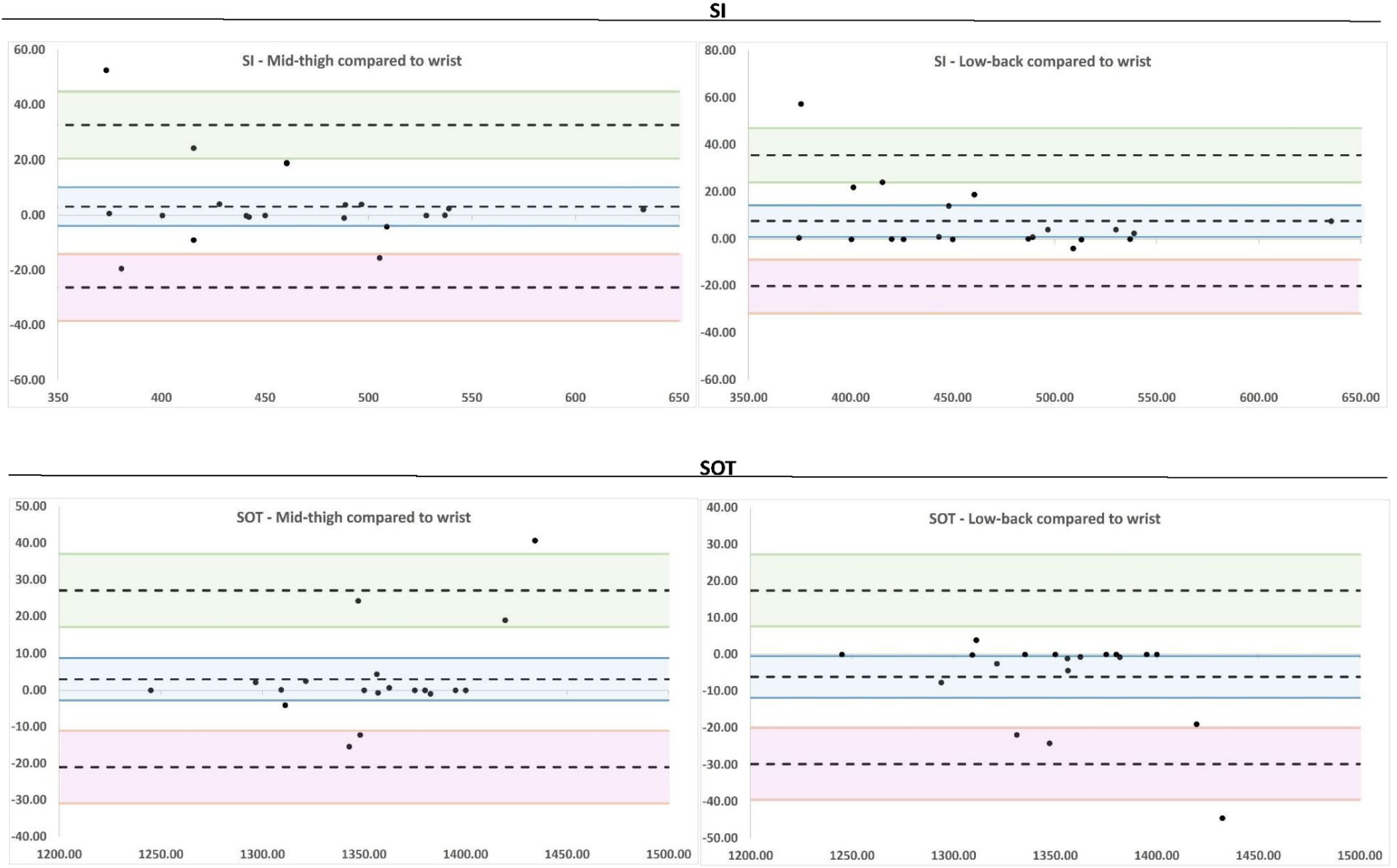
Bland-Altman plots showing the individual differences between the mid-thigh and wrist, and low-back and wrist locations for each of the sleep metrics. Each participant Is identified as a dot. The middle-dotted line represents the mean difference. The upper and lower dotted lines represent the 95% limits of agreement. The shaded areas represent the confidence interval limits. *Plot unit scales may differ

**Table 4.** Results of Bland-Altman and Pearson Correlation analysis comparing sleep metrics obtained from low-back and wrist device placements.

| N=20 | Low-back compared to wrist |  |  |  |  |  | Pearson correlation, r | p value | 95% CI |
| --- | --- | --- | --- | --- | --- | --- | --- | --- | --- |
|  | Bias | 95% CI | Lower LoA | 95% CI | Upper LoA | 95% CI |  |  |  |
| TST, min | 22.50 | 11.50, 33.40 | -23.30 | -42.3, -4.32 | 68.3 | 49.30, 87.28 | 0.92 | p < 0.001 | 0.82, 0.97 |
| WASO, min | -14.80 | -23.53, -60 | -51.50 | -66.75, -36.26 | 22.00 | 6.71, 37.20 | 0.81 | p < 0.001 | 0.56, 0.92 |
| SE, % | 3.28 | 1.42, 5.14 | -4.51 | -7.74, -1.27 | 11.08 | 7.84, 14.31 | 0.78 | p < 0.001 | 0.51, 0.91 |
| AWK, n | -3.45 | -5.85, -1.05 | -13.49 | -17.66, -9.32 | 6.59 | 2.42, 10.76 | 0.63 | p < 0.003 | 0.26, 0.84 |
| SI, min | 7.70 | 1.04, 14.36 | -20.19 | -31.77, -8.62 | 35.59 | 24.02, 47.17 | 0.98 | p < 0.001 | 0.95, 0.99 |
| SOT difference, min | -6.15 | -11.79, -0.51 | -29.75 | -39.55, -19.96 | 17.45 | 7.66, 27.25 | 0.97 | p < 0.001 | 0.92, 0.99 |

Abbreviation: CI, confidence interval
LoA, Limits of Agreement

participants across both mid-thigh and low-back comparisons to the wrist. Not surprisingly three of these four were identified in the visual inspection as mentioned above. Scrutiny of these four showed large and/or frequent periods of awake time between intraparticipant devices. There was, however, no consistency with how the devices were out of sync with each other on either an intra or interparticipant comparison of the minute-by-minute visual representations of their sleep. A single explanation of cause across all four outlying participants is likely unrealistic. However, some explanations that could posited for the various participants could be lack of protocol compliance, device detachment and reapplication, excessive restlessness, or a (movement to a) sleeping position that did not meet the requirements and threshold for the van Hees algorithm to register sleep. Further detailed examination of the data collected from these participants would be required to identify true causality of the discrepancies.

From a clinical perspective, the wide limits of agreement suggest that individual estimates from the thigh may not be suitable for identifying individual-level changes in sleep metrics. However, the narrow 95% CI of the bias estimate suggest that the sample mean of sleep metrics from the thigh and wrist devices are in good agreement, except for AWK, which is slightly underestimated by the thigh unit. The precision of the bias estimates and LoA may improve with larger sample sizes.

The back-mounted device significantly overestimated TST, SE, and SI, and significantly underestimated WASO, AWK, and SOT compared to the wrist-mounted device (Table 5). The widest LoA was observed for TST (-23.3 min to 68.3 min; 91.6 min). Similarly wide LoA are observed for all sleep metrics which suggests that the back-mounted device does not produce comparable estimates to the wrist-mounted device.

**Table 5.** Cohen’s d of the mid-thigh and low-back devices.

|  | Wrist |  | Mid-thigh |  | **d** | Low-back |  | **d** |
| --- | --- | --- | --- | --- | --- | --- | --- | --- |
|  | mean | standard deviation | mean | standard deviation |  | mean | standard deviation |  |
| TST, min | 403.10 | 60.62 | 410.01 | 59.99 | 0.11 | 425.57 | 59.26 | 0.37 |
| WASO, min | 60.57 | 31.53 | 56.87 | 31.59 | -0.12 | 45.80 | 26.46 | -0.51 |
| SE, % | 0.87 | 0.06 | 0.88 | 0.06 | 0.14 | 0.90 | 0.05 | 0.56 |
| AWK, n | 16.95 | 5.72 | 14.55 | 5.73 | -0.42 | 13.50 | 6.13 | -0.58 |
| SI, min | 463.68 | 68.54 | 466.88 | 65.14 | 0.05 | 471.38 | 63.11 | 0.12 |
| SOT difference, min | 1357.15 | 47.37 | 1354.11 | 42.15 | 0.07 | 1351.00 | 43.06 | -0.14 |

Cohen’s d values demonstrated small effect size in the comparison of mid-thigh to wrist devices. The exception to this was AWK (d= 0.42).

Low-back Cohen’s d values were consistently larger than those of the mid-thigh groups. WASO, SE, and AWK all showed a moderate effect size.

G*Power (Version 3.1.9.7; Faul et al., 2020) was used for the post-hoc analysis to assess the statistical power of the study, revealing that the sample size was sufficient. The results of the F-tests, specifically ANOVA with repeated measures indicated a high likelihood of correctly rejecting the null hypothesis at 99% (β = 0.993). Moreover, the t-tests utilising the correlation point biserial model demonstrated a range of 93% to 100% likelihood of correctly rejecting the null hypothesis for the lowest and highest correlations observed (β = 0.929 and β = 1.00, respectively).

## Discussion

The present study investigated whether an open-source sleep algorithm designed for use with wrist-based accelerometers could produce similar sleep metric estimates from accelerometric data collected from either a mid-thigh or lower-back placement. This study aimed to contribute to the advancement of accurate and comprehensive sleep tracking methodologies that can be used for sleep focused, and/or 24-hour time use studies. The key findings of this study indicate that sleep metrics obtained from the van Hees algorithm (2015) using a mid-thigh placement are better than those from a low-back placement and comparable to those from a wrist placement. Building on previous work, an immediate impact of this is that this method can now be used in retrospective data analysis, offering both increased value to participants and stakeholders of the research.

Biases of under- and overestimation of accelerometery devices with regards to sleep quantity and quality metrics are an acknowledged and unresolved aspect of their use in sleep tracking. Incidences of under and overestimation vary from study to study (Johansson et al., 2022; Tonetti et al., 2008; Toon, 2016; van Hees et al., 2015) and no standardised device placement, algorithm, or settings setup has shown itself to emerge as the field standard of excellence (Evenson et al., 2022). Other groups have emphasized the development of tracking methods that utilize thigh data to advance the field (Stevens et al., 2020). Johansson et al. (2022) have also taken steps towards the development of a thigh-based sleep tracking algorithm. Their sleep metric results contrasted those of the present study in that TST, SE, and SI were respectively under-rather than overestimated. These comparisons can only be superficial however as the baseline comparison to each of these respective studies is different. TST, SE and SI estimates generated by the thigh (and back) device placements of this study align more consistently with those of the van Hees et al. (2015) study where these metrics were overestimated. Both the Johansson et al. (2022) and van Hees et al. (2015) study used PSG as their ground truth in training their respective algorithms. A natural progression for the continuation of inquiry for this work would be to take this algorithm and device location into a comparison against PSG itself to identify where further biases may lay.

This research also has the potential to reduce researcher and participant burden in computing sleep metrics. Previous research such as Tudor-Locke (2014) involved researchers manually identifying sleep onset, wake and total sleep times (using accelerometery from the hip). The statistical evidence of the present study having such a close relationship to its baseline dataset indicates that this method is sufficient in accuracy for the measurement of sleep on a group level in free-living settings. Further, the sample frequency and sensitivity rate of the present study also coincide with the default sampling frequency settings of 100Hz in AX3 devices creating further ease of use and potential applicability to previously collected data sets (Narayanan et al., 2020; Stewart et al., 2018). This may also be effective for other brands of accelerometery devices as Plekhanova et al. (2020) found that when analysed identically, sleep outcomes of different devices were comparable across studies at a group level. Participant burden can potentially be reduced in future studies by requiring fewer accelerometers for comprehensive 24-hour time use behaviour analysis. Using a dual accelerometer system on the mid-thigh and low-back has been shown to be an effective way of measuring posture and movement in free-living environments (Narayanan et al., 2020; Stewart et al., 2018). The results of this study indicate that the thigh mounted accelerometery can also be used as a non-inferior method of sleep metric estimation thus eliminating the need for a wrist mounted device that is unable to detect body positioning (Rosenberger, 2019).

One of the future directions this study prompts is its potential to integrate sleep positioning identification and duration into sleep metrics and by extension into the research within the field of 24-hour time use. Unlike traditional methods, which can neglect data about (sleep) posture, the results of this study suggest an ability to leverage data collected from the mid-thigh that can not only be used in analysing sleep but can also extrapolate more robust knowledge around the interplay of body positioning and position duration. However, for the true positioning of body postures, the positional orientation of more than a single limb is likely required. In this vein, the retention of a device placed on the low-back would likely greatly improve positional identification (Narayanan et al., 2020).

## Limitations

There are some limitations that must be acknowledged with the results of this study. Firstly, while the power of this study is strong, the sample size of this study is not on a large scale. As well, while a free-living sleep environment does allow the participant a familiar sleep location for data collection, it also opens the study up to variables that cannot necessarily be controlled for between participants such as partners in the same bed causing movement, pets, or children that could all cause changes to a participants’ body angles and movement patterns.

Additionally, the results of this study do not comprehensively remove the need for sleep diaries. The van Hees algorithm has been updated to function without the need for a sleep diary guider (van Hees et al., 2018). Further analysis would need to be completed to identify if results were still consistent without the use of the sleep diaries but is worthy of further investigation.

## Conclusions

This study shows that there is scope for mid-thigh accelerometery to be evaluated for sleep metrics using the van Hees algorithm when wrist accelerometery is not available. The study also highlights the need for increased availability of open-source measurement tools for interpreting data collected using accelerometers mounted on the thigh and lower back to assess activity over time domains that include sleep periods.

## Acknowledgements

The authors would like to thank Tom Stewart for help with the data processing.

## Author Contributions

**Geoff Passfield**: conceptualisation, investigation, data collection, methodology, formal analysis, visualisation, writing – original draft, writing – review and editing, project administration.

**Lisa Mackay**: conceptualisation, methodology, writing – review and editing, supervision, resources.

**Catherine Crofts**: methodology, writing – review and editing, formal analysis, supervision.

**Grant Schofield**: supervision, writing – review and editing.

**Funding Information**: This research received no specific grant from any funding agency, commercial or not-for-profit sectors.

## Competing Interests

The authors declare none.

## Data availability statement

The data that support the findings of this study are available from the corresponding author upon reasonable request.

## Ethics approval statement

The authors assert that all procedures contributing to this work comply with the ethical standards of the relevant national and institutional committees on human experimentation and with the Helsinki Declaration of 1975, as revised in 2008.

## Notes

### Competing Interest Statement

The authors have declared no competing interest.

### Funding Statement

This study did not receive any funding

### Author Declarations

Ethical approval was obtained from Auckland University of Technology ethics committee (AUTEC 22/300).

